# Intrauterine contraceptive device-related perceptions and perspectives among community members and healthcare providers in southern Ethiopia: An exploratory descriptive qualitative study

**DOI:** 10.1101/2025.07.17.25331520

**Authors:** Samson Dubale, Mihiret Tesfaw

## Abstract

**Background:** Despite a notable incremental trend in the use of contraceptive methods, the use of intrauterine contraceptive devices (IUCDs) in Ethiopia is still less than 2%. This study aims to provide an in-depth exploration of the perceptions held by community groups, IUCD users and health care providers to support efforts of addressing the existing gap in service uptake.

**Methods:** Exploratory descriptive qualitative (EDQ) research design was conducted between September 1 and October 1, 2024, in Sodo town. A maximum variation sample of 23 study participants was recruited using a purposive sampling technique. A total of 16 in-depth interviews and 7 key informant interviews were audio-taped and transcribed verbatim. A semistructured in-depth interview guide and a field book were used. Thematic analysis was conducted via Colaizzi seven-step data analysis framework, and the open code software version 4.03 was utilized for data analysis and management.

**Results:** Five main themes and 12 subthemes emerged from the study. The identified themes are poor knowledge of IUCD, male partners’ involvement in decision-making, perceptions and perspectives on IUCD, users’ experience and realities, and optimizing IUCD service uptake. Findings from in this study revealed that the use of the IUCD method is not solely dependent on the choice of women.

**Conclusion:** The acceptance and utilization of IUCD service is strongly affected by misconceptions within both the community and healthcare providers. Targeted awareness initiatives and gender-inclusive literacy programs are vital to enhance service uptake.

**What is already known on this topic?:** Despite increasing contraceptive use in Ethiopia, IUCD uptake remains low, hindered by myths, male partner influence, and health system barriers.

**What this study adds?:** This study provides nuanced insights into community and provider perspectives, highlighting the need for gender-inclusive contraceptive literacy.

**How this study might affect research, practice, or policy?:** Findings advocate for targeted awareness campaigns addressing the deep-rooted misconceptions and male partners’ engagement, informing policies to improve IUCD access and equity in Ethiopia’s family planning programs. Applying novel designs will encourage researchers to focus on recruiting study participants through the maximum variation sampling technique, enabling understanding of the phenomena of interest from different point of view.

## Introduction

Intrauterine contraceptive devices (IUCDs) are a type of modern contraceptive option for women and girls, which are small T-shaped devices made of plastic and sometimes contain copper or hormones. It is one of the most widely used reversible contraceptive methods globally, with only 8 pregnancies per 1000 women in the first year of use for copper-bearing IUDs and 2 per 1000 women for levonorgestrel-releasing IUDs or LNG-IUDs. The copper-bearing IUD is effective for 10–12 years, and the LNGIUD is effective for 5 years, both with immediate fertility returns once removed[1].

Despite a notable increase in the overall contraceptive prevalence in Ethiopia, the utilization of the IUCD is still at its lowest level. According to the latest EDHS report of Ethiopia, IUCD use among women in the reproductive age group is very low, accounting for 1.4% at the country level and 1.3% in the South Nation, Nationalities, and Peoples (SNNP) region of the country[2].

The underutilization of the IUCD in Ethiopia can be attributed to various factors, ranging from different community-based perceptions toward the IUCD, myths and misconceptions, and health system-related barriers. The unfriendly nature of health care facilities and providers, including their perceptions, biases, and knowledge and skill gaps regarding IUCDs, can greatly affect the quality of counseling and service provision. Research in Ethiopia revealed that IUCD method usage is also strongly dependent on husbands/partners’ involvement, women’s literacy status, women’s perceptions of the method, and the availability of a consistent and credible source of information about the IUCD[3], [4], [5].

This study therefore, aims to provide an in-depth exploration of the perceptions held by community groups, IUCD users and health care providers. The insights gathered will inform targeted policy interventions to improve IUCD service uptake, ultimately contributing to better sexual and reproductive health outcomes for women and girls in Ethiopia.

## Methods

The findings of this study followed the COREQ checklist, which is a consolidated criteria for reporting qualitative research[6]. (Supplementary file 2)

### Study Setting and Design

An exploratory descriptive qualitative research[7] was conducted between September 01 and October 01, 2024 in Sodo town, southern Ethiopia. Wolaita Sodo city is the administrative capital of the Wolaita Zone, located 328 kilometers south of Addis Ababa. On the basis of the 2019 population projection by the CSA of Ethiopia, Sodo town has a total population of 137,522 people[8].

### Sampling and Participant Recruitment Procedure

A sample of 23 study participants from the community and healthcare setup was purposefully selected under the three health center catchments of Sodo town. Study participants, including IUCD users, non-IUCD users, husbands/partners of women currently using the IUCD method, health care providers working in family planning rooms, and community-based health extension workers (HEWs), were included based on their specific characteristics relevant to the aim of the study. This participant recruitment procedure aims to ensure a diversified sample from both the community and the health facility setup that helps gain deep insight into the phenomenon of interest. The purpose of the study and the scope of its involvement are explained to the study participants. (Supplementary material-1 section) The guidance of community-based health extension workers (HEWs) and the medical records of contraceptive users at those facilities were important sources of information.

A maximum variation purposive sampling technique[9] was employed to select the 23 study participants. Sixteen in-depth interviews (IDIs) and seven key informant interviews (KIIs) were conducted. The decision on the number of study participants considers previous literature experiences, recommendations of research books, and idea saturation (the occurrence of redundant information)[10], [11], [12]. Study participants’ recruitment is presented in Table 1.

**Table 1:** Sampling distribution of the study Participants.

| SN | Catchment Facilities | Study Participants |  |  |  |  | Total |
| --- | --- | --- | --- | --- | --- | --- | --- |
|  |  | IUCD User's | Non-IUCD contraceptive users | Partners of IUCD Users' | HCPs | HEWs |  |
| 1 | Geneme HC catchment | 2 | 1 | 2 | 2 | 1 | 8 |
| 2 | Sodo HC catchment | 2 | 2 | 2 | 1 | 1 | 8 |
| 3 | Wadu HC catchment | 2 | 1 | 2 | 1 | 1 | 7 |
| 4 | Total | 6 | 4 | 6 | 4 | 3 | 23 |

### Data collection tools and procedures

A semistructured open-ended IDI guide (Supplementary material-1 section a) and KII guides (Supplementary material-1 section b), supplemented by a field notebook, were used to collect data from the respondents. The tool was tested on 2 participants before the actual interview. The primary investigators of the study (males, senior expert and a PhD student) who had a clear understanding of the ethical and technical aspects of the qualitative research, collected rich data. The interviews were conducted at the health facility and participants’ homes in convenient private areas. An interview protocol was followed to maintain the logical flow of the IDI sessions[13]. All interviews were audio-taped and supportive field notes were also taken during every interview session. On average, the interviews lasted for 24 minutes.

### Data quality management and analysis

**R**epeated listening to the Amharic version of the audio recorded data was conducted to understand the message from each participant. The audio transcripts were then changed to Amharic (local language) text transcripts and then to English by an expert in both languages. The transcripts and field notes were prepared in rich text file format for import into Open Code software. Open code software version 4.03 was used for data coding and management. The seven steps of Colaizzi’s inductive phenomenological data analysis framework were thoroughly followed to analyze qualitative data and gain valuable insights into the experiences and perspectives of the study participants[14]. The analysis runs simultaneously with the data collection process. It follows an inductive way of formulating significant statements from transcripts, developing categories from codes, and then developing themes from categories/subthemes. Finally, findings from this study are presented in the form of themes, subthemes, and richly descriptive participant quotes.

The current study maintained the five principles of trustworthiness, including credibility, transferability/applicability, dependability, conformability, and authenticity [15] to maintain its rigor.

## Results

### Socio-demographic information of the study participants

A total of twenty-three individuals from various backgrounds were included in this study. The participants’ ages ranged from 20--48 years, resulting in a mean age of 30.4 years. Among the total participants, 26% (6) were current IUCD users, 17.4% (4) were non-IUCD method users, 26% (6) were partners of IUCD users, 17.4% (4) were healthcare providers, and the remaining 13% 93) were community-based HEWs. Most of the participants, 14 (60.8%), were females, and the remaining 9 (29.2%) were males. All participants lived in urban areas, and most of them belonged to the protestant religion (56.5%), Orthodox Christians (30.4%), Muslims (8.7%), and Catholic followers (the other 4.3%). (Table 2)

**Table 2:** Background characteristics of the study participants.

| S. No | Participant's Code | Sex | Marital Status | Educational status | Occupation | Participant Type/Background |
| --- | --- | --- | --- | --- | --- | --- |
| 1 | IDIP-01 | Female | Married | BSC Degree | Employed | IUCD user |
| 2 | IDIP-02 | Female | Married | Secondary school | Housewife | IUCD user |
| 3 | IDIP-03 | Female | Married | Primary School | Merchant | IUCD user |
| 4 | IDIP-04 | Female | Married | Primary School | Housewife | IUCD user |
| 5 | IDIP-05 | Female | Married | BSC degree | Employed | IUCD user |
| 6 | IDIP-06 | Female | Married | Unable to read and write | Daily Laborer | IUCD user |
| 7 | IDIP-07 | Female | Married | Primary school | Student | Non-IUCD method user |
| 8 | IDIP-08 | Female | Single | BSC Degree | Employed | Non-IUCD method user |
| 9 | IDIP-09 | Female | Divorced | Secondary School | Unemployed | Non-IUCD method user |
| 10 | IDIP-10 | Female | Married | Primary school | Housewife | Non-IUCD method user |
| 11 | IDIP-11 | Male | Married | Primary school | Merchant | IUCD users' Partner |
| 12 | IDIP-12 | Male | Married | BSC Degree | Employed | IUCD users' Partner |
| 13 | IDIP-13 | Male | Married | BSC Degree | Construction | IUCD users' Partner |
| 14 | IDIP-14 | Male | Married | Unable to read and write | Private job | IUCD users' Partner |
| 15 | IDIP-15 | Male | Married | BSC degree | Merchant | IUCD users' Partner |
| 16 | IDIP-16 | Male | Married | Primary | Merchant | IUCD users' Partner |
| 17 | KIIP-01 | Female | Married | College Diploma | Employed | Community-based HEW |
| 18 | KIIP-02 | Female | Married | College Diploma | Employed | Community-based HEW |
| 19 | KIIP-03 | Female | Single | College Diploma | Employed | Community-based HEW |
| 20 | KIIP-04 | Male | Single | BSC degree | Employed | FP Provider |
| 21 | KIIP-05 | Female | Single | BSC degree | Employed | FP Provider |
| 22 | KIIP-06 | Male | Single | BSC degree | Employed | FP Provider |
| 23 | KIIP-07 | Male | Married | BSC degree | Employed | FP Provider |

### Themes Emerged in the Study

Five themes, twelve subthemes, and 98 codes emerged from this EDQ study. The identified themes are mentioned as follows. (Figure 1)

**Figure 1:**
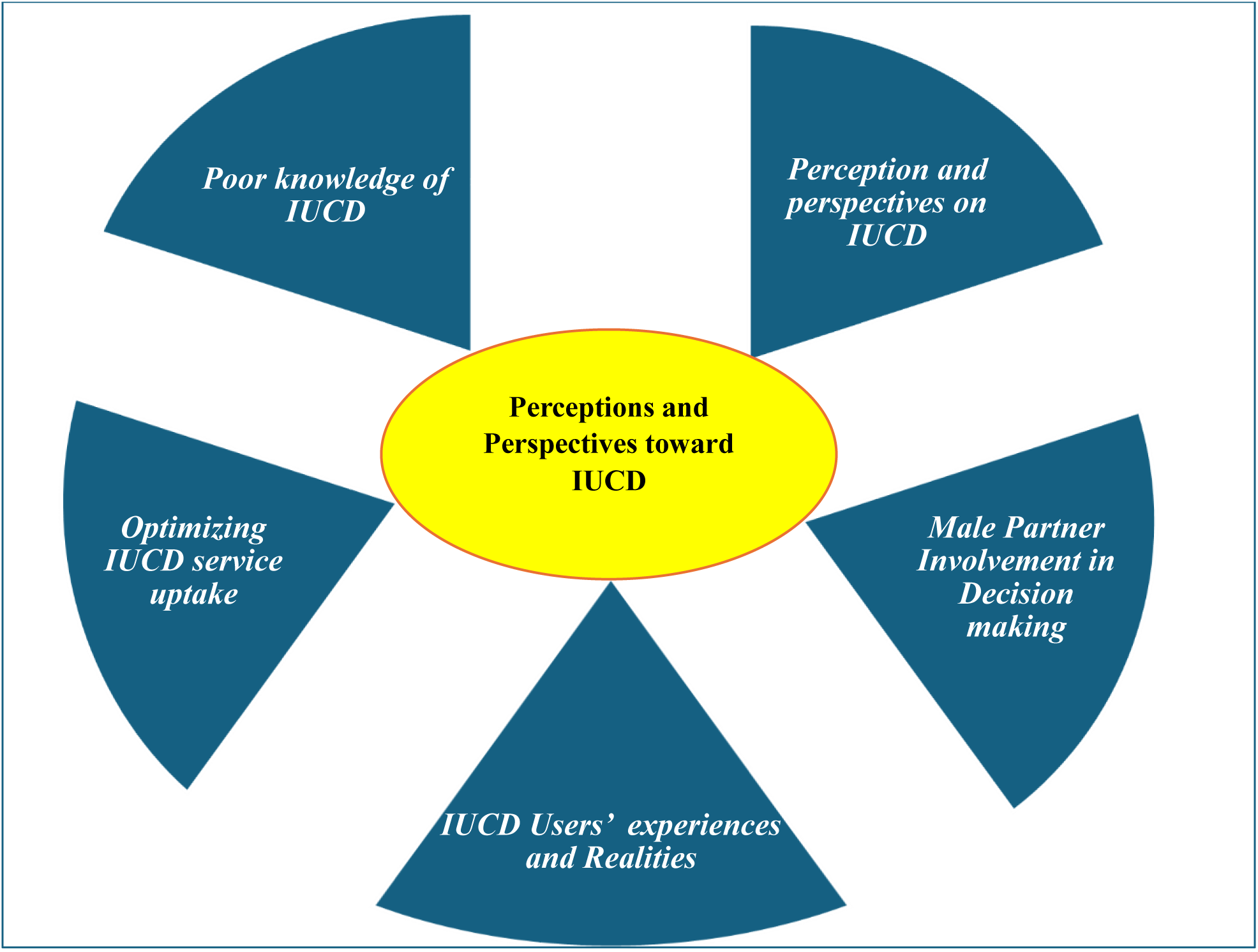
Illustrates the major themes emerged from the study

#### Theme 1: Poor knowledge of IUCD

##### Subtheme 1: Community Awareness

In the present study, some study participants had minimal understanding of the IUCD method, and even a few of them had misconceptions regarding it. One of the IDI participants discussed that,

> “I don’t know about it. (She seems confused and frustrated to talk.) …umm… truly speaking, I have no idea about this type. I think there is no such option at our nearby facilities … maybe it will be available in bigger cities, the healthcare providers here told me about the injectable option only, and I used it for the last 5 years.” (IDIP-07, non-IUCD method user)

##### Subtheme 2: Health care providers’ communication

Many participants revealed that the role of healthcare providers in providing detailed information regarding what the IUCD method is, how it works, its uses, side effects, and possible risks has been overlooked. Owing to a lack of adequate information and tailored counseling, clients are using short-term methods of contraception. One participant said that,

> “… The last time I took the injectable method, the providers told me to change it with a new method, something which will be kept in the uterus, and I refused to take it on the spot because I didn’t have any clue about it previously”. (IDIP-10, non-IUCD method user)

##### Subtheme 3-Myths and Misconceptions

This study explores the presence of multiple rumors and misconceptions regarding the IUCD method circulating within the community. Some of the misconceptions are that the IUCD causes infertility, migrates to the head and other body parts, and causes cancer, among others. One of the IDI participants explained,

> “I heard that it causes infertility and has discomfort during sexual intercourse, and there are many rumors about IUCD in our community.” (IDIP-15, IUCD user’s partner)

#### Theme 2: Male Partners’ Involvement in Decision Making

##### Subtheme 1: Lack of Partner Approval

Unlike the supportive husbands, some of the study participants complain about the controlling and restrictive behavior of their husbands, which leads them to frustration and even to use the service secretly. One of the study participants from IDI explained:

> “My husband didn’t know about this issue. If he knew that he would attack me, I decided not to tell him about my current family planning use. Sometimes he asks me why I failed to become pregnant and argues with me to remove the previously inserted implant. However, I am responding that I already removed the implant, and if it is God’s wish, you need to wait.” (IDIP-03, IUCD user)

##### Subtheme 2: Availability of Partner Support

The presence of partner/husband support is identified as an encouraging situation for women to choose the IUCD method. One participant mentioned his involvement in decision-making as

> “I support her in any way, including her nutritional status and others. I believe that our children should be raised and grow properly. Therefore, I told my wife that if we continue having children, this will highly affect our economy and future. For this reason, we need to discuss spacing childbearing by using long span methods such as a loop. Then, she agreed with my idea and took the service.” (IDIP-11, partner of the IUCD user)

#### Theme 3: Perceptions and Perspectives on IUCD

##### Subtheme 1: Mothers’ perceptions of IUCD

The reason for the negative perceptions mentioned by most of the study participants is related to the existing knowledge and information gap in the method and the presence of community-wide rumors, myths, and misconceptions attached to the IUCD.

An IDI participant said that,

> “Umm…Honestly speaking, I do not have a good view of this method of birth control (to say IUCD) because I have heard many complaints and side effects from my neighbors. Most people said that if you take this method before having your first baby, it will cause a delay in pregnancy when you want to have a baby, it will cause infertility, and the like. Owing to this information that I have from the community, I have a negative view of the method.” (IDIP-09, non-IUCD method user)

##### Subtheme 2: Provider‒client interaction

Most of the KII participants in this study revealed that IUCD is the most effective nonhormonal type of contraceptive option for women with space childbearing. A community-based HEW said that:

> “Honestly speaking, I have many reservations about recommending loop (to say IUCD) methods for my clients. Many clients are worried about the insertion pain and its side effects, because these thoughts that can discourage them from considering this option, it’s hard to change their minds at my level.” (KII-01, community-based HEW)

##### Subtheme 3: Sociocultural influences

Community-level negative observations toward the IUCD also contributed to the limited utilization of the method in the study area. The community-based HEWs mentioned that the widespread community-based sociocultural contributing factors affected the use of this method of contraception.

> “As a community-based health extension worker during my home-to-home visit, I frequently noticed that many of the husbands of IUCD-user women believe that using the IUCD method causes discomfort while having sexual intercourse with their partners.” (KIIP-03, Community-based HEW)

#### Theme 4: IUCD Users’ Experience and Realities Subtheme 1: Reasons for choosing the IUCD method

The main reason for choosing the IUCD for most of the study participants was fear of the side effects of other contraceptive methods and its non-hormonal nature. One of the study participants explained as follows:

> “Yeah… umm… The reason for me to use this option (referring to the IUCD) is related to my own lifestyle and working conditions.” (IDIP-03, IUCD user)

#### Theme 2: Mother’s journey with the method

Most of the study participants using the IUCD method are navigating their contraceptive choices, highlighting the importance of voluntary informed decision-making, healthcare providers’ support, and empowerment in obtaining the service. One of the participants expressed this as follows:

> “Ene ye IUCD adinaki negn” (an Amharic phrase meaning I am a fan of the IUCD method). The reason why I am supporting IUCD use is that it is free from any pain during insertion … and … also no need to remember daily or monthly once it is inserted correctly.” (IDIP-01, IUCD user)

#### Theme 5: Optimizing IUCD Service Uptake

##### Subtheme 1: Fostering Community Awareness

Improving community awareness of the IUCD service is the major strategy used by most of the service providers who have participated in the study. This includes husband involvement, organizing information, education, and communication activities at colleges and universities. One of the service providers described this as follows:

> “It is better if we provide the service at the community level in outreach, integrating with health extension workers, I believe we will improve the service uptake. Most of the mothers who come from rural areas will face transportation problems, so if we provide the service there in the community, we will serve more clients.” (KIIP-07, FP provider)

##### Subtheme 2: Health System-Related Intervention

In addition to awareness creation activities, participants emphasized the need for continuous capacity building for health service providers, as the service requires adequate skills and competency. The presence of an adequate number of trained providers on the IUCD can improve service provision. One of the study participants was described as follows:

> “Many providers trained on the IUCD are not currently providing the IUCD service, mainly due to skill atrophy. I think a continuous mechanism of professional development needs to be in place to equip family planning service providers’ skills and knowledge so that the service uptake will improve.” (KIIP-05, FP provider)

## Discussion

The study revealed that there is a limited understanding of the IUCD method as a contraceptive option among participants. A lack of knowledge about the IUCD method is prevalent among non-IUCD method users, as evidenced by a comparatively better understanding and utilization of the injectable type of contraceptive options than the IUCD. This finding is in line with results obtained from a previous mixed-method study conducted in Ethiopia[17]. The latest national demographic and health survey report of the country also has similar findings to the current identified theme that more than 50% of women do not know the IUCD method[2].

Most study participants revealed that there is limited supportive involvement of male partners, even those who are unaware of their wives’ current IUCD use. These findings are consistent with several previous studies conducted in Ethiopia[17], [19], [20], [21]. In contrast to the findings of a previous study from Nepal that revealed that all women using the IUCD method had been discussed with their partners[22], the current findings revealed that some women are using the IUCD method without disclosing to their partners. This emphasizes the need for meaningful engagement of male partners at all levels of sexual and reproductive health decision-making, addressing their concerns, enhancing shared responsibility, and ultimately contributing to improved IUCD service uptake[23].

While some IDI and KII participants recognized its safety and effectiveness, others held reservations about the side effects, inserted places, and doubted the presence of pain during sexual intercourse. This aligns with existing evidence that explores the complex interplaying nature of determinant factors for the limited service uptake of IUCD, informing the necessity of tailored interventions at both the community and health facility levels[4], [10], [17], [24].

In contrast to a previous study from the Angacha district of southern Ethiopia[25], which reported that 19% of IUCD methods use discontinuation within a year, findings from this study revealed that all women used the method for at least two years, indicating that the discontinuation/premature removal of the IUCD service is not a problem once they start using it. Despite the low utilization in the study area [26], the current findings reveal that the IUCD is an effective and efficient form of contraception that improves the work and productivity of users and enhances their families’ health and well-being. This finding is in line with studies conducted in Ethiopia[24], Tanzania[27], and Indonesia[28].

Furthermore, the involvement of male partners and satisfied clients during health education sessions enhances women’s acceptance and utilization of the IUCD method. These findings are consistent with current working national reproductive health strategies and the WHO’s recommendations[16],[29],[30].

## Limitation of the Study

Even though this research meets its objectives, all the participants in the study were from an urban setting, and the study did not address the perspectives and experiences of rural dwellers.

## Conclusion

The acceptance and utilization of IUCD service is strongly affected by misconceptions within both the community and healthcare providers. Targeted awareness initiatives and gender-inclusive literacy programs are vital to enhance service uptake. Programs aimed at engaging satisfied IUCD-users can also help to dispel fears and misunderstandings related to the IUCD methods.

## Data availability statement

The data generated in this study are not publicly available due to the sensitivity of the research subject and the ethical importance of protecting the anonymity of the study participants but are available from the corresponding author upon reasonable request.

## Availability of competing interests

The authors declare that they have no competing interests.

## Patient and public involvement

Patients and/or the public were not involved in the design, conduct, reporting, or dissemination plans of this research.

## Ethics Approval

The study was approved by the institutional review board of Wolaita Sodo University with a reference number of CHSM/ERC/07/21. Written informed consent is obtained from study participants after providing.

## Author contributions

Both authors (SD and MT) contributed equally to the conception, drafting, analysis, and interpretation of findings in this study. The authors of this work also conducted a critical review of the findings and finally agreed on the journal to which the article is submitted.

## Data Availability

All data produced in the present study are available upon reasonable request to the authors.

## Acknowledgments

Our deepest gratitude goes to the study participants who devotedly shared their insights into the phenomena of interest in this study. We are also thankful to the zonal health department, the town health office, the three health facility leadership teams, and the providers for their cooperation in conducting this study.

## Author’s Information

### Author and Affiliations

**SD:** Field operations deputy director, MSI Ethiopia Reproductive Choices, Hawassa, Ethiopia.

**MT:** Mary Joy Ethiopia, Quality Improvement Specialist, USAID FFHPCT Activity, Hawassa, Ethiopia, PHD candidate at Wolaita Sodo University, Department of Public Health, Sodo, Ethiopia

