## Supplemental material 1 for "Intrauterine contraceptive device-related perceptions and perspectives among community members and healthcare providers in southern Ethiopia: An exploratory descriptive qualitative study"

I. **Background Information of the IDI Participants**

Please, tell me about yourself.

1. Age ----------
2. Sex -----------
3. Marital status -----------
4. Educational level ----------
5. Residence ---------------
6. Religion-----------------------
7. Occupation------------
8. Participant background (IUCD User, non-IUCD method user, partners of IUCD users, family planning providers, community-based health extension workers)
II. **IDI Guide: Open-ended questions to explore IUCD-related Perceptions and Perspectives among the community members**
1. What did you know about IUCD (loop)?

- How did you learn about IUCD? Source of information?

2. What are your thoughts on using an IUCD?
3. What advantages and disadvantages do you associate with IUCD?
4. Have you used an IUCD method before?

- If yes, please describe your detailed experience in using the IUCD method.
- If not, what factors influenced you not to use this method?

5. How do cultural beliefs affect the IUCD use?
6. What types of myths and misconceptions do you hear about IUCD in your community?

- What do you think about these rumors?

7. How did you describe your interaction with healthcare providers while discussing IUCD-related issues? Please discuss in detail:

- Any barriers
- Enablers

8. Have you faced any challenges in accessing IUCD services? Please describe its nature and occurrence in detail.
9. What recommendations do you suggest to improve IUCD acceptance and utilization in your community in the future?

**III. Closing**

1. What else would you like to be clarified? If there is anything more you would like to add, please continue.

Thank you for participating. This has been a very successful discussion. Your opinions will be an asset to the study and to improvement efforts. I hope you have found the discussion interesting. If there is anything you are unhappy with or wish to complain about, please contact or speak to me later. I would like to remind you that any comments featured in this report will be anonymous.

### **‘Supplementary Material’ Section 2: Key Informant Interview Guide**

**Intrauterine contraceptive device-related perceptions and perspectives among community members and healthcare providers in southern Ethiopia: An exploratory descriptive qualitative study Background Information of the KII Participants**

Please, tell me about yourself.

1. Age----------
2. Sex-----------
3. Marital status-----------
4. Educational level----------
5. Residence ---------------
6. Religion-----------------------
7. Occupation------------
8. Participant background (IUCD User, non-IUCD method user, partners of IUCD users, family planning providers, community-based health extension workers)
I. **KII Guide: Open-ended questions to explore IUCD-related Perceptions and Perspectives among healthcare providers and community HEWs**
1. How do you describe IUCD as a method of contraception?
2. How do you describe the general understanding of women and girls about the IUCD method?
3. How did people in your locality obtain information regarding the IUCD method?
4. What are the common attitudes and perceptions you encountered regarding IUCD? Please describe in detail.
5. Have you heard any myths and misconceptions regarding the IUCD method?
6. What cultural barriers to IUCD usage do you know in this community?

- How did such cultural barriers affect women’s contraception choices?
- What are the health facilities' measures?

7. What is the health facilities' responsiveness to IUCD service in terms of:

- Information exchange with clients and the public
- Access and availability of IUCD supplies and services
- Training and capacity-building activities

8. What major challenges do women encounter in accessing IUCD services at your facilities and in this community?
9. What strategies do you recommend to improve IUCD service uptake in your catchment areas?
II. **Closing**
1. What else would you like to be clarified? If there is anything more you would like to add, please continue.

Thank you for participating. This has been a very successful discussion. Your opinions will be an asset to the study and improvement efforts. I hope you have found the discussion interesting. If there is anything you are unhappy with or wish to complain about, please contact or speak to me later. I want to remind you that any comments featured in this report will be anonymous.

#### **‘Supplementary Material-1 Section c: participant information sheet and consent form**

**Intrauterine contraceptive device-related perceptions and perspectives among community members and healthcare providers in southern Ethiopia: An exploratory descriptive qualitative study**

We want to thank you for taking the time to meet with us today. We would like to know your experiences and perspectives on the IUCD method of contraception. We are exploring women’s experience and perception towards the bilateral tubal ligation service. The interview should take less than an hour. We will be tape recording the session to avoid losing your comments. All responses will be kept confidential. This means that your interview response will only be shared with research team members, and we ensure that no one can identify your response. You are free not to talk about anything you don’t want to, and you might end the interview anytime.

I have read the participant information sheet, and I have been given opportunities to raise questions, and all of my questions about the study have been answered. I understand that the IDI will be recorded. I also accept that the anonymized data I gave may be published in a journal article.

I agree to participate in this study.

Interviewer name -------------------------- signature ---------------------- Date------------------------

Interviewee code-------------------------- Signature----------------------- Date------------------------
